# Pulse-wave harmonic signatures of cardiovascular regulatory coherence to a cognitive challenge in mild cognitive impairment: a community-based cross-sectional study

**DOI:** 10.64898/2026.04.24.26351647

**Authors:** Kun-Yuan Hsiao

## Abstract

To evaluate whether hemodynamic responses to acute cognitive stress, measured via pulse-wave harmonic analysis, can characterize cardiovascular regulatory coherence and differentiate older adults with mild cognitive impairment (MCI) from cognitively intact individuals, this exploratory cross-sectional observational study utilized a within-session pre–post cognitive task design. A total of 101 community-dwelling older adults in southern Taiwan were stratified by Montreal Cognitive Assessment (MoCA) scores into Reference (MoCA ≥26; n=12, paired n=10), MCI (MoCA 18–25; n=50, paired n=45), and dementia-level (MoCA <18; n=39) groups, the latter being excluded from task-evoked analyses. The primary outcome was the Harmonic Response Consistency Score (HRCS), quantifying the directional uniformity of cardiovascular regulatory responses, alongside secondary measures of harmonic amplitudes (Cn) and phase angles (Pn). Although mean pre–post changes were subtle, response organization differed by cognitive status. The Reference group exhibited high response consistency (mean HRCS = 9.00), characterized by coordinated harmonic down-modulation. Conversely, the MCI group showed attenuated, directionally heterogeneous responses. Compared to the Reference group, the MCI group demonstrated significantly lower HRCS values for the Cn domain (Mean difference: 2.60, 95% CI 0.29–4.91; p=0.020) and PnSD domain (Mean difference: 1.98, 95% CI 0.04–3.92; p=0.030), indicating a breakdown in regulatory coherence. These findings suggest that acute cognitive stimulus reveals coherent harmonic down-modulation in cognitively intact older adults but fragmented responses in MCI. Pulse-harmonic profiling thus serves as a robust physiological index of cardiovascular regulatory coherence, which, when integrated with neuropsychological assessments, may enhance the sensitivity of non-invasive, community-based screening frameworks for early cognitive aging.

## Introduction

Cognitive decline and dementia are increasingly conceptualized not solely as disorders of brain function but as systemic conditions accompanied by widespread physiological alterations [1–3]. In older adults, cognitive impairment frequently coexists with dysregulation of cardiovascular, metabolic, and autonomic systems [4–6]. These co-occurring changes suggest that cognitive decline reflects an integrated physiological state rather than an isolated neurological deficit. Such systemic characteristics are particularly evident in community-dwelling and rural older populations, where chronic disease burden and age-related physiological vulnerability tend to accumulate [7].

Within this systemic framework, numerous studies have reported associations between cognitive impairment and health-related indicators, including blood pressure, glycemic control, lipid profiles, body composition, and emotional status. Anxiety, depressive symptoms, metabolic dysregulation, and vascular burden commonly accompany cognitive decline in later life [5, 6]. Importantly, a recent systematic review integrating evidence across multiple cognitive domains demonstrated that metabolic syndrome is not uniformly associated with global cognition, but shows domain-specific relationships with memory, executive function, and processing speed [8]. Large-scale cohort evidence further indicates that metabolic syndrome is associated with poorer cognitive performance and increased dementia risk in community-dwelling populations [9]. However, these health-related measures are often examined independently, and their inter-relationships are rarely integrated into a unified physiological description of cognitive decline, limiting the ability to characterize it as a coherent physiological phenotype in real-world community settings.

Pulse wave signals provide a compact representation of cardiovascular dynamics that inherently integrate cardiac output, vascular properties, and autonomic regulation. These regulatory systems are closely linked to metabolic health and cognitive function. At the population level, metabolic syndrome has been shown to be associated with increased arterial stiffness, a vascular property that directly influences pulse wave morphology and propagation characteristics [10]. Accordingly, pulse wave characteristics may serve as a composite physiological signature of the systemic state accompanying cognitive decline. Importantly, pulse wave measurement is non-invasive and feasible in community-based contexts, enabling physiological characterization outside specialized clinical facilities [11, 12].

Harmonic analysis of pulse waves enables a structured frequency-domain characterization of waveform morphology, stability, and temporal organization [13, 14]. Harmonic amplitudes, variability, and phase-related indices capture complementary aspects of cardiovascular regulation across cardiac cycles [15–17]. Rather than acting as isolated diagnostic markers, these features describe underlying physiological patterns that may reflect regulatory integrity or dysregulation, making them suitable for exploratory investigation of heterogeneous conditions such as cognitive decline.

Physiological systems are inherently dynamic and responsive to contextual demands. More recent work using photoplethysmography has demonstrated that pulse wave amplitude dynamically tracks cognitive load during task performance, supporting the sensitivity of pulse-derived measures to cognitive engagement [18, 19]. Examining how pulse wave patterns are modulated—or remain inflexible—during cognitive engagement may therefore provide insight into regulatory flexibility associated with cognitive decline. In this context, cognitive tasks serve as standardized perturbations to probe short-term regulatory dynamics, rather than as diagnostic tests.

Beyond the magnitude of task-evoked changes, the internal coherence of cardiovascular responses across harmonic orders may provide a complementary indicator of regulatory integrity.

In complex biological systems, adaptive regulation is characterized not only by responsiveness, but by coordinated, directionally consistent adjustments across multiple scales. Conversely, early dysregulation may manifest as fragmented or contradictory responses, even when mean group-level changes appear small.

The selection of a mental arithmetic task as a stressor is grounded in its well-established capacity to elicit centrally mediated autonomic arousal. Serial subtraction requires sustained working memory and executive control, processes that activate the cortico-limbic network, particularly the prefrontal and anterior cingulate cortices [20, 21]. These brain regions are integral components of the central autonomic network (CAN), which regulates the peripheral cardiovascular system via the vagus and sympathetic nerves [22]. In healthy physiology, this acute sympathy-excitation typically induces peripheral vasoconstriction and transient arterial stiffening, which theoretically manifests specific alterations in the pulse waveform morphology—characteristically, a reduction in the amplitude of harmonics [17]. Consistent with this, task-related reductions in peripheral pulse amplitude have been reported in PPG-based cognitive load paradigms [19]. However, this adaptive chain of events relies on the integrity of the neuro-cardiovascular axis. It was hypothesized that in individuals with early cognitive decline, the coupling between central cognitive effort and peripheral vascular response might be compromised [23, 24], resulting in blunted or aberrant pulse wave modulation compared to the coherent vasoconstrictive response expected in cognitively intact individuals [25–27].

In this community-based study, pulse-wave harmonic analysis was used to characterize cardiovascular waveform structure and beat-to-beat variability across cognitive strata defined by MoCA. The primary aim was to describe within-subject changes in harmonic features immediately before and after a subtraction task as a standardized cognitive perturbation. Secondary aims were to examine cross-sectional associations between baseline harmonic parameters and health-related measures (blood pressure, anthropometrics, and laboratory markers), and to explore phase-related metrics given their expected lower sensitivity to short cognitive engagement. In addition to examining pre–post changes in individual harmonic features, the study aimed to characterize the consistency of task-evoked responses across the harmonic spectrum (C1–C10) as a potential marker of regulatory coherence in cognitive aging. This study aimed to characterize task-evoked pulse harmonic responses and their coherence across cognitive strata in a community-dwelling older population.

## Materials and methods

### Study Design and Setting

This observational, community-based study examined associations between pulse harmonic features, health status, and cognitive function in older adults. A within-subject pre–post protocol was used to assess short-term pulse-wave responses immediately before and after a subtraction task. The within-subject pre–post design was used to reduce inter-individual confounding, though residual confounding by age and education cannot be excluded. This was a cross-sectional observational study with a within-session pre-post physiological assessment. All assessments were completed in a single session. Participants were recruited from community healthcare settings in the Cishan and Meinong areas (Kaohsiung City, Taiwan), and the recruitment period for this study was from 01/04/2025 to 30/10/2025. The reporting of this study adheres to the STROBE guidelines for cross-sectional studies; the full STROBE checklist is available as Supporting Information.

### Participants

Community-dwelling adults aged ≥65 years were recruited. Participants were required to complete pulse-wave measurements and basic cognitive assessments. A total of 101 participants were enrolled (16 men, 85 women; mean age 78.8 ± 6.1 years). Exclusion criteria included acute unstable medical conditions; known neurological disorders or psychiatric conditions that could substantially affect autonomic/cardiovascular regulation or hinder participation; and any conditions likely to interfere with study procedures or compromise pulse-wave recording quality. MoCA-based groups were used for descriptive stratification rather than clinical diagnosis. Participants with MoCA < 18 were retained in the cross-sectional cohort description but were excluded from task-evoked analyses due to inability to reliably complete the cognitive task or missing post-task recordings.

The study was approved by the Institutional Review Board of National Cheng Kung University Hospital (B-ER-114-044). Written informed consent was obtained from all participants, including consent for questionnaire administration, physiological measurements, and authorized access to hospital electronic medical records.

#### Patient and Public Involvement

Patients and the public were not involved in the design, reporting, or dissemination plans of this research. However, local community hubs and volunteers assisted in the conduct of the study by promoting the research activities and contacting potential participants to attend the assessment sessions.

### Measures and Procedures

#### Health and Cognitive Assessments

Demographic data and anthropometric measures (height, weight, waist circumference) were obtained by interview and on-site measurement. Cognitive function was assessed using the Montreal Cognitive Assessment (MoCA), cognitive complaints using the AD8, and emotional status using the Beck Anxiety Inventory (BAI) and Beck Depression Inventory (BDI).

Resting systolic and diastolic blood pressure were measured using an automated monitor. Biochemical indicators (fasting glucose, total cholesterol, triglycerides, and high-density lipoprotein cholesterol) were retrieved from hospital electronic medical records after participant consent. All assessments were completed prior to pulse-wave recording.

#### Pulse-Wave Measurement

Pulse-wave signals were recorded using a photoplethysmography-based system (Biosignalsplux Solo Kit; PLUX Wireless Biosignals S.A., Portugal) after a minimum of five minutes of seated rest in a quiet, temperature-controlled environment. Signals were sampled at 1000 Hz with a resolution of up to 16-bit using finger-mounted sensors. Each recording lasted 3 minutes to obtain a sufficient number of consecutive pulse beats for robust beat-level harmonic estimation while maintaining participant tolerance. To minimize measurement bias, pulse-wave recordings were standardized with fixed sensor placement, sampling rate, and recording duration, and analyses focused on within-subject pre–post changes.

Pulse-wave recordings were obtained three times within a single session: an initial baseline recording during quiet seated rest to allow signal stabilization and participant acclimatization, a pre-task (pre-cognitive) recording obtained immediately before the subtraction task, and a post-task (post-cognitive) recording obtained immediately after task completion. The primary task-evoked physiological change (Δ) was defined as the difference between post-cognitive and pre-cognitive recordings, while the baseline recording was used for signal quality verification and descriptive purposes. The procedure is shown in Fig 1.

**Fig 1.**
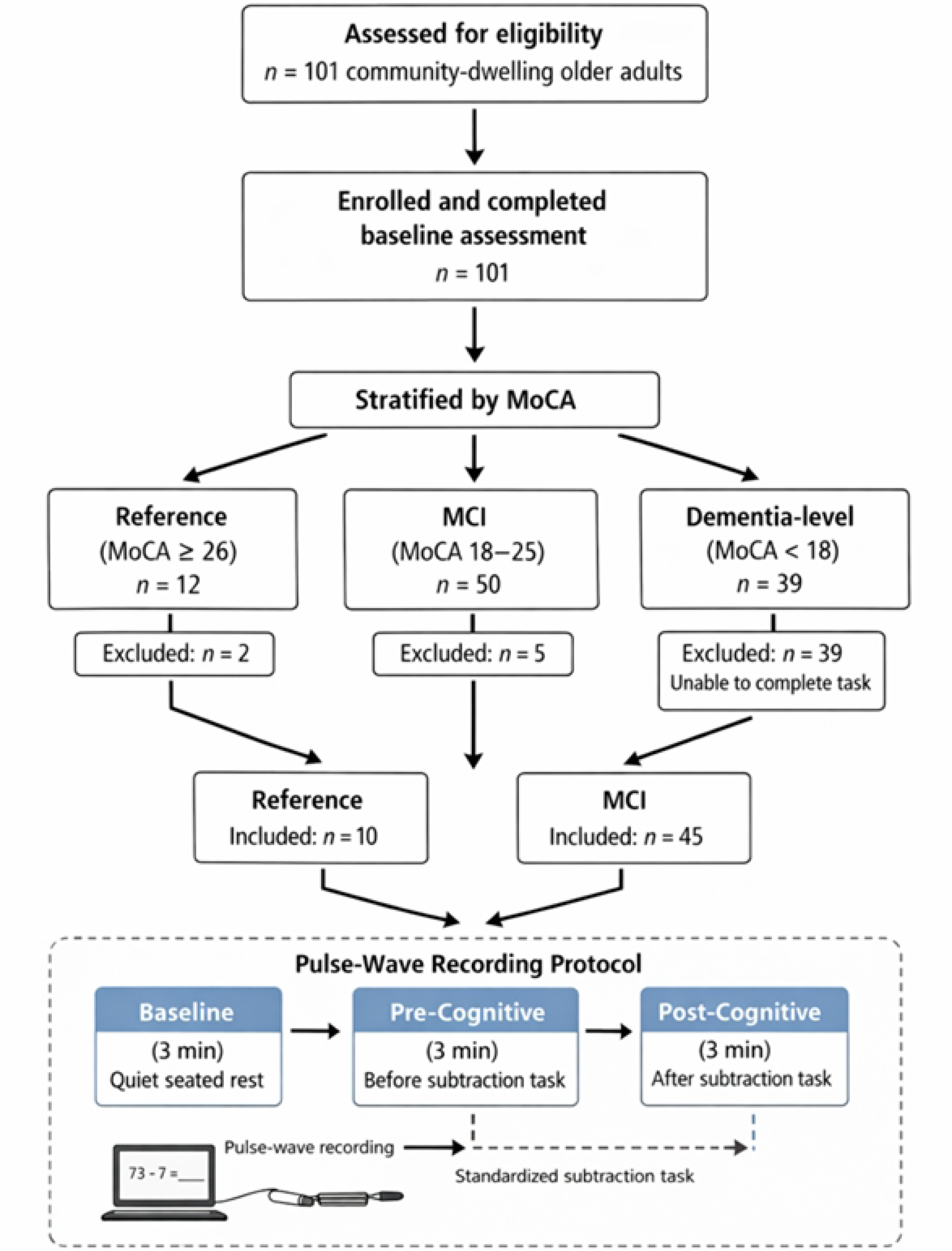
Study flow and pulse-wave recording protocol. Flow of participants from recruitment to final paired analysis and schematic overview of the pulse-wave recording protocol. After MoCA-based stratification and exclusion of dementia-level participants and recordings failing quality control, 55 participants (10 Reference; 45 MCI) were included in paired analyses. Pulse-wave signals were recorded at baseline, immediately before, and immediately after a subtraction task, and task-evoked changes were defined as post minus pre recordings.

Flow of participants from recruitment to final paired analysis and schematic overview of the pulse-wave recording protocol. After MoCA-based stratification and exclusion of dementia-level participants and recordings failing quality control, 55 participants (10 Reference; 45 MCI) were included in paired analyses. Pulse-wave signals were recorded at baseline, immediately before, and immediately after a subtraction task, and task-evoked changes were defined as post minus pre recordings.

#### Subtraction Task

Participants completed a computerized subtraction task consisting of 40 consecutive true/false items. For each item, participants judged whether the displayed subtraction equation was correct or incorrect using a binary response. Pulse-wave signals were recorded for 3 minutes immediately before (pre-cog) and immediately after (post-cog) task completion. Task accuracy was recorded for descriptive analyses.

### Experimental Protocol

Participants completed interviews, questionnaires, and cognitive assessments, followed by a pulse-wave recording after sitting rest. Pulse-wave signals were then recorded for 3 minutes immediately before the subtraction task (pre-task) and for 3 minutes immediately after task completion (post-task). This procedure enabled within-subject characterization of short-term changes in pulse harmonic features associated with cognitive task engagement. Participants were included in paired analyses only if both pre-cog and post-cog recordings were available and passed quality control. Pulse harmonic parameters (Cn, Pn, CVn, and PnSD) were summarized for baseline, pre-cog, and post-cog recordings, whereas task-evoked responses were quantified using Δ = post − pre.

### Signal Processing and Statistical Analysis

#### Pulse Harmonic Analysis

Pulse-wave signals were processed using MATLAB. Raw signals were visually inspected, and segments with excessive motion artifacts were excluded prior to analysis. Fast Fourier transform (FFT) was applied at the single-beat level, with each pulse waveform segmented on a beat-by-beat basis and transformed individually from the time domain into the frequency domain. Harmonic components were extracted from each individual pulse waveform and subsequently summarized across beats within each recording segment, including:

. Cn: the normalized amplitude of the n-th harmonic component relative to the fundamental frequency, representing the strength of specific harmonic oscillations. Cn was computed as the amplitude of the n-th harmonic normalized to the fundamental component (An/A1).

. Pn: the phase angle of the n-th harmonic component, reflecting temporal shifts in harmonic structure. Phase angles were analyzed descriptively without applying circular statistical transformations.

. CVn: the coefficient of variation of the n-th harmonic amplitude across pulse cycles, representing beat-to-beat variability.

. PnSD: the standard deviation of the phase angle of the n-th harmonic component, reflecting phase instability across cycles.

These harmonic parameters have been widely applied in cardiovascular and integrative medicine research and summarized in prior systematic reviews [17].

### Statistical Analysis

Analyses were performed using MATLAB and JASP (version 0.18.3.0). Continuous variables are presented as mean ± standard deviation. Normality was assessed using the Shapiro–Wilk test. For each participant, task-evoked change scores were computed as Δ = post − pre for each parameter. Pre–post changes were evaluated using paired-sample t-tests for normally distributed variables or Wilcoxon signed-rank tests otherwise. Participants with missing or poor-quality pre- or post-task recordings were excluded from paired analyses. Given the exploratory aim, all harmonic orders within each parameter family (Cn, Pn, CVn, and PnSD) were analyzed using the same procedure. Associations between baseline pulse harmonic parameters and clinical variables (MoCA, AD8, BAI, BDI, anthropometrics, blood pressure, and biochemical indicators) were examined using Pearson correlations. Tests were two-tailed with p < 0.05. Given the exploratory objective, while emphasizing effect direction, variability/heterogeneity, and cross-harmonic consistency across harmonic orders; results are interpreted as descriptive patterns rather than confirmatory evidence.

To quantify the regulatory coherence of the cardiovascular system across the frequency spectrum, the Harmonic Response Consistency Score (HRCS) was calculated for four physiological domains: Cn, CVn, Pn, and PnSD. The HRCS is not intended as a diagnostic classifier, but as a summary index quantifying the degree of directional coherence across harmonic orders in response to cognitive perturbation. For each domain, the task-evoked change of the first ten harmonics was calculated (post minus pre) and binarized, assigning a value of +1 for an increase and −1 for a decrease. These directional values were summed to produce a raw score, and the absolute value (ranging from 0 to 10) was derived to index the magnitude of coherence independent of direction; a score of 10 indicates uniform regulation, while a score of 0 reflects a fragmented response. Because the score is based on ten binary (+1/−1) directions, absolute HRCS values take even-numbered steps (0, 2,…, 10). Inter-group differences in absolute HRCS values between the Ref. and MCI groups were primarily analyzed using the Mann-Whitney U test due to the unequal sample sizes; however, independent samples t-tests were also computed for reference to verify the robustness of the statistical findings. Because the expected direction of task-evoked change may vary across individuals and baseline states, the analyses focused on directional uniformity (coherence) rather than assuming a single canonical direction.

## Results

### Participant Characteristics

A total of 101 community-dwelling older adults were included in the community health dataset (16 males, 85 females; mean age 78.8 ± 6.1 years). Of the 101 enrolled participants, 55 were included in paired pre–post analyses after excluding those with missing or low-quality recordings. Most participants had ≤ elementary school education, and chronic cardiometabolic conditions were common (hypertension, diabetes, and dyslipidemia). Based on MoCA scores, participants were categorized into three groups: the Reference (Ref.) group (MoCA ≥ 26), the MCI group (MoCA 18–25), and the Dementia group (MoCA < 18). These terms denote MoCA-based cognitive strata rather than confirmed clinical diagnoses. Hereafter, Ref. and MCI refer to MoCA-based cognitive strata used for descriptive purposes. Table 1 summarizes demographic characteristics, education, disease burden, questionnaire scores (MoCA, AD8, BAI, BDI), anthropometrics, blood pressure, and laboratory indices for the study cohort.

**Table 1.** Demographic and clinical characteristics of participants stratified by cognitive status.

| Characteristic | Ref.<br>(MoCA $\geq 26$) | MCI<br>(MoCA 18–25) | Dementia<br>(MoCA $< 18$) | P value |
| --- | --- | --- | --- | --- |
| N | 12 | 50 | 39 | — |
| Female, n (%) | 10 (83.3%) | 40 (80.0%) | 34 (87.2%) | 0.71 |
| Male, n (%) | 2 (16.7%) | 10 (20.0%) | 5 (12.8%) |  |
| Age (years) | $74.75 \pm 5.88$ | $78.26 \pm 6.14$ | $83.56 \pm 5.04$ | $<0.001$ |
| Education level | | | | $<0.001$ |
| —Uneducated | 0 (0.0%) | 9 (18.0%) | 17 (43.6%) |  |
| —Elementary school | 4 (33.3%) | 25 (50.0%) | 19 (48.7%) |  |
| —Junior high school | 2 (16.7%) | 13 (26.0%) | 2 (5.1%) |  |
| —Senior high school | 3 (25.0%) | 2 (4.0%) | 0 (0.0%) |  |
| — Junior college | 2 (16.7%) | 0 (0.0%) | 0 (0.0%) |  |
| — College | 1 (8.3%) | 0 (0.0%) | 0 (0.0%) |  |
| — University | 0 (0.0%) | 1 (2.0%) | 0 (0.0%) |  |
| **MoCA score** | 27.33 ± 0.49 | 21.68 ± 2.31 | 13.92 ± 3.34 | <0.001 |
| **AD8 score** | 1.00 ± 0.85 | 1.68 ± 2.37 | 3.26 ± 4.72 | <0.001 |
| **BAI score** | 1.83 ± 1.64 | 3.72 ± 3.80 | 4.23 ± 3.64 | 0.01 |
| **BDI-II score** | 2.33 ± 1.61 | 4.04 ± 4.30 | 4.41 ± 4.41 | 0.03 |
| **Height (cm)** | 158.58 ± 7.63 | 156.99 ± 6.56 | 155.41 ± 6.36 | 0.18 |
| **Weight (kg)** | 59.86 ± 8.78 | 58.83 ± 9.23 | 58.98 ± 8.55 | 0.92 |
| **BMI (kg/m<sup>2</sup>)** | 23.83 ± 2.92 | 23.86 ± 3.69 | 24.42 ± 3.32 | 0.64 |
| **Waist circumference (cm)** | 82.25 ± 7.90 | 83.38 ± 10.56 | 82.34 ± 8.10 | 0.83 |
| **Systolic BP (mmHg)** | 141.92 ± 21.57 | 136.18 ± 16.68 | 130.74 ± 17.70 | 0.04 |
| **Diastolic BP (mmHg)** | 73.08 ± 15.63 | 67.55 ± 10.77 | 64.21 ± 9.19 | 0.02 |
| **Fasting glucose (mg/dL)** | 139.67 ± 53.37 | 127.17 ± 43.63 | 132.80 ± 55.07 | 0.58 |
| **Total cholesterol (mg/dL)** | 187.78 ± 22.09 | 192.43 ± 33.50 | 190.32 ± 34.63 | 0.81 |
| **Triglycerides (mg/dL)** | 100.00 ± 22.98 | 107.20 ± 43.89 | 109.00 ± 44.46 | 0.76 |
| **HDL cholesterol (mg/dL)** | 62.00 ± 9.90 | 62.50 ± 14.28 | 60.40 ± 14.90 | 0.88 |

Values are presented as mean ± standard deviation or n (%). Cognitive status was classified based on Montreal Cognitive Assessment (MoCA) scores as follows: Reference stratum (MoCA ≥ 26), mild cognitive impairment (MCI; MoCA 18–25), and dementia-level impairment (MoCA < 18). P values were calculated using one-way analysis of variance (ANOVA) for continuous variables and chi-square tests for categorical variables, as appropriate. For task-evoked analyses, paired sample sizes were reduced to Ref. (n = 10) and MCI (n = 45) due to data quality control.

### Task-evoked pulse harmonic responses (pre–post)

Task-evoked pulse responses were evaluated using within-subject pre–post comparisons of pulse-wave recordings obtained immediately before and immediately after the subtraction task. Participants in the dementia group were excluded from the task-evoked analysis because they were unable to complete the subtraction task, resulting in missing post-task recordings. In addition, datasets with unrecoverable technical errors were excluded. After excluding participants with corrupted pre- or post-condition recordings (which precluded within-subject pairing), the final paired sample sizes for the subtraction-task analyses were 10 participants in the Ref. group and 45 participants with MCI. In the overall cohort, the Ref. stratum included 12 participants (MoCA ≥ 26); two were not included in paired analyses because either the pre- or post-task recording was missing or failed quality control.

#### Subtraction Task: Harmonic Amplitudes (Cn)

Overall, task-evoked changes in harmonic amplitudes were small in magnitude across harmonic orders. In the Ref. subgroup, harmonic amplitudes showed a generally consistent decrease following the subtraction task, although none of the pre–post changes reached statistical significance in paired-sample tests (all p > 0.05). In the MCI subgroup, mean changes in harmonic amplitudes were close to zero and exhibited mixed directions across harmonics, with no component reaching statistical significance (all p > 0.05). Pre–post changes in harmonic amplitudes (ΔCn) for the Ref. and MCI subgroups are illustrated in Fig 2.

**Fig 2.**
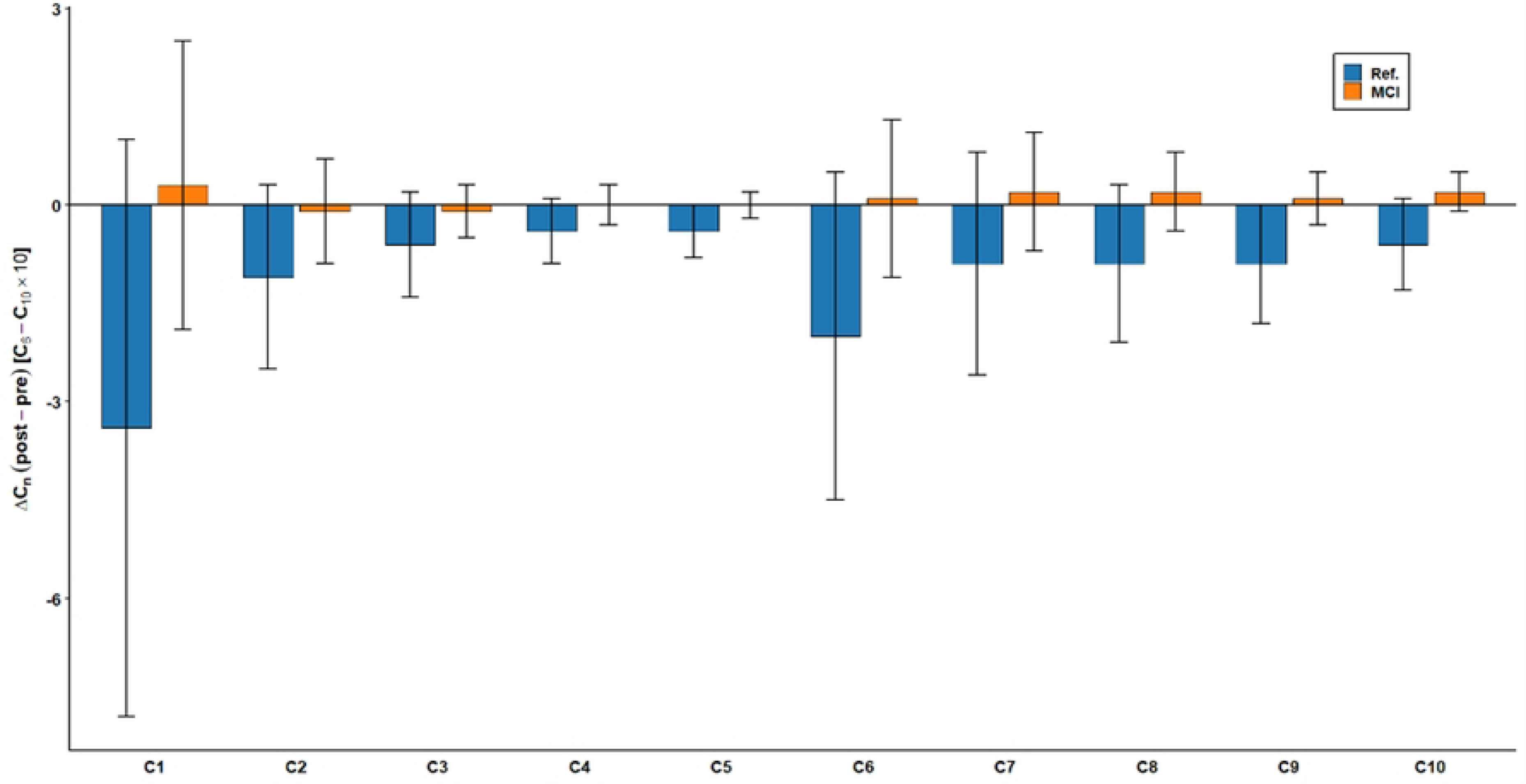
Task-evoked changes in harmonic amplitudes (ΔCn) following the subtraction task, stratified by cognitive status. Mean task-evoked changes in pulse harmonic amplitudes (**Δ**Cn = post-task − pre-task) across harmonic orders (C1–C10) are shown for participants in the Reference (Ref.) group and those with mild cognitive impairment (MCI). Points represent group means, and error bars indicate standard errors (SE). For enhanced visualization of higher-order trends, values for **Δ**C6–**Δ**C10 were scaled by a factor of 10; however, all formal statistical analyses were performed using unscaled raw values to ensure scientific rigor.

#### Subtraction Task: Variability Indices (CVn)

Pre–post changes in beat-to-beat variability indices (CVn) were generally small across harmonic orders. In both the Ref. and MCI subgroups, mean ΔCVn values showed mixed directions and did not reach statistical significance in paired comparisons (all p > 0.05). Error bars indicated substantial inter-individual variability, particularly for lower-order harmonics, but no consistent task-evoked pattern was observed across CVn measures. Pre–post changes in CVn are summarized in Fig 3.

**Fig 3.**
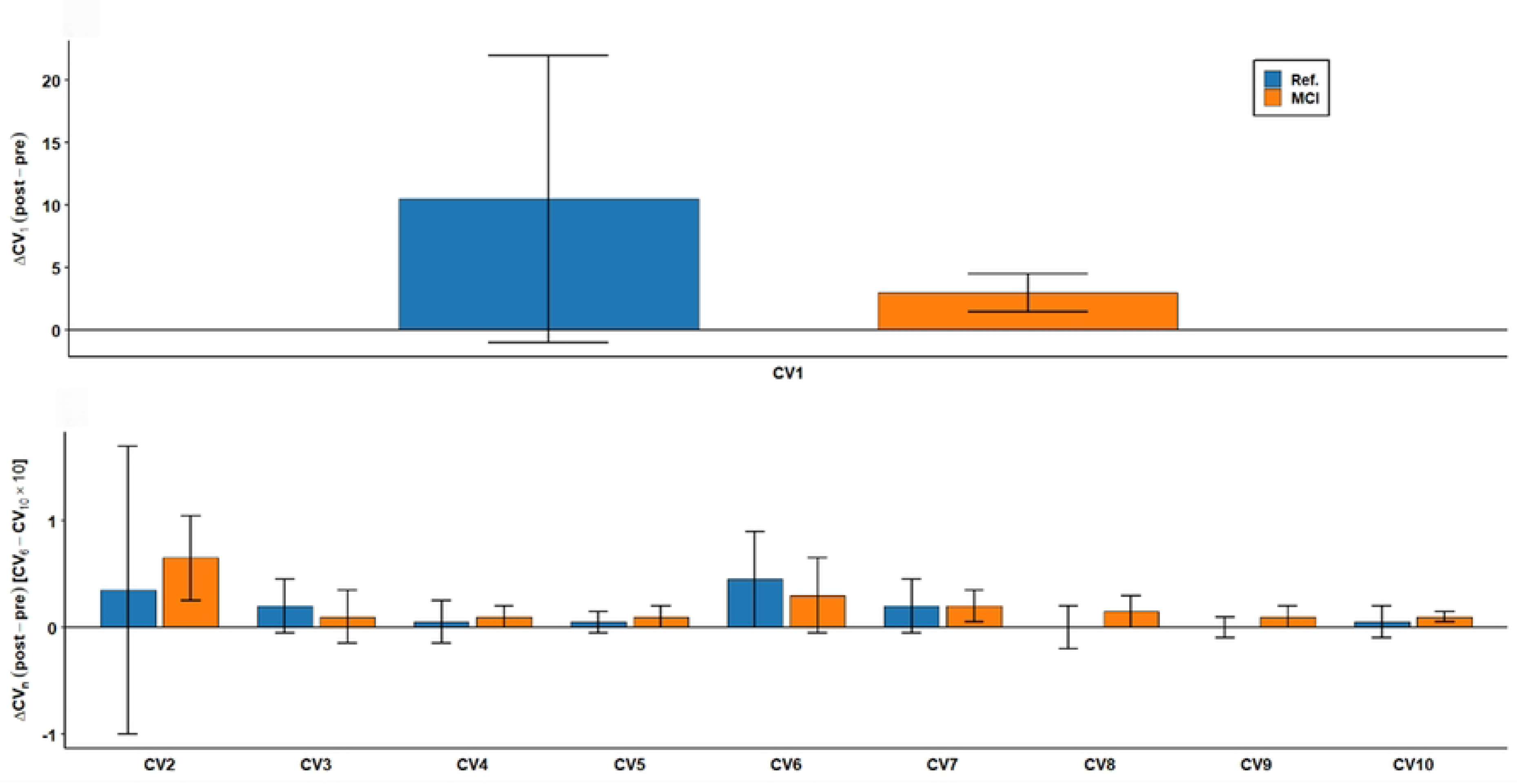
Task-evoked changes in variability indices (ΔCVn) following the subtraction task, stratified by cognitive status. Mean task-evoked changes in coefficients of variation (ΔCVn = post-task − pre-task) across harmonic orders are shown for the Reference (Ref.) and mild cognitive impairment (MCI) groups. Points represent group means, and error bars indicate standard errors (SE). The upper panel displays ΔCV1 separately due to its larger dispersion, while the lower panel presents ΔCV2–ΔCV10 on a dedicated scale to facilitate comparison among higher-order harmonics.

#### Subtraction Task: Phase-Related Measures (Pn and PnSD)

Phase-related measures demonstrated limited sensitivity to the subtraction task, with overall small pre–post shifts observed across harmonic orders. In the Ref. subgroup, phase parameters (P1–P10) remained largely stable, and none of the paired comparisons reached statistical significance. In the MCI subgroup, P1 and P2 showed nominally significant pre–post changes (uncorrected p < 0.05), whereas higher-order phase components were non-significant. Given the exploratory nature of the analyses and the number of comparisons performed, these findings should be interpreted cautiously. Phase variability, indexed by PnSD, showed small pre–post shifts across harmonics in both subgroups, and no PnSD parameter reached statistical significance. Pre–post changes in phase-related measures (Pn and PnSD) across harmonic orders are presented in Fig 4 and Fig 5.

**Fig 4.**
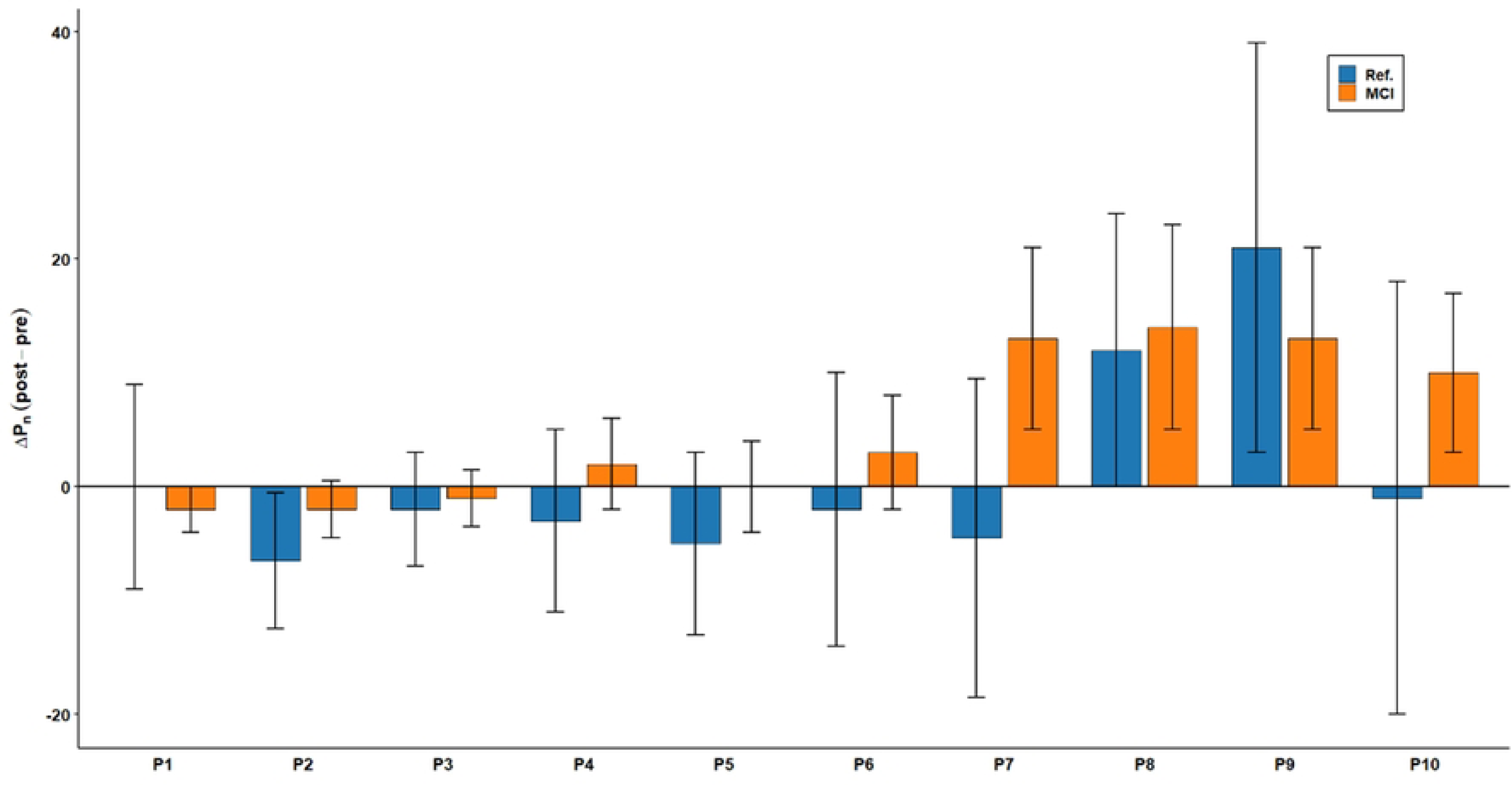
Task-evoked changes in phase parameters (ΔPn) following the subtraction task, stratified by cognitive status. Mean task-evoked changes in phase angles (ΔPn = post-task − pre-task) across harmonic orders (P1–P10) are shown for the Reference (Ref.) and mild cognitive impairment (MCI) groups. Points represent group means, and error bars indicate standard errors (SE). Asterisks indicate nominal statistical significance for within-group pre–post comparisons (*p < 0.05).

**Fig 5.**
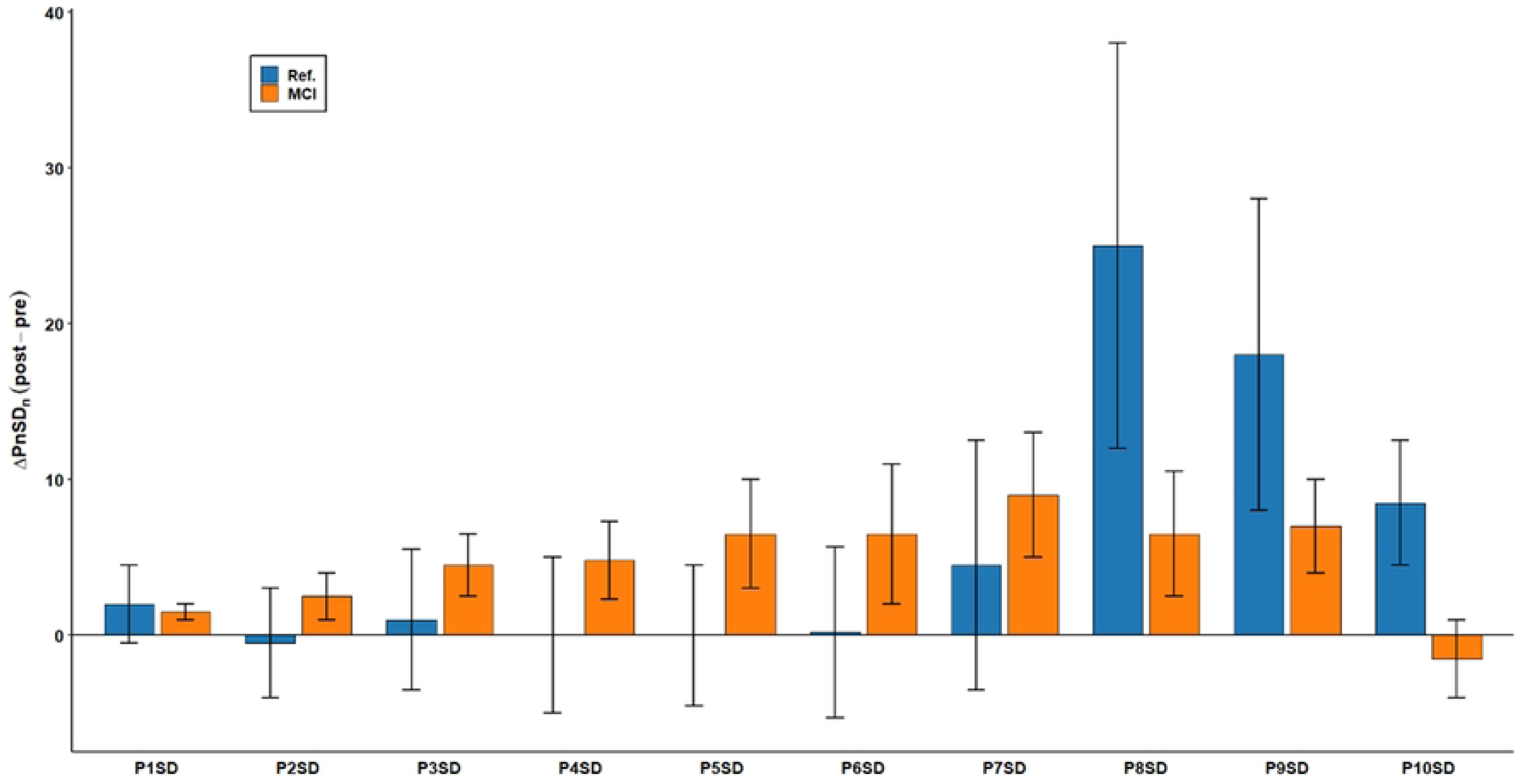
Task-evoked changes in phase noise standard deviation (ΔPnSD) following the subtraction task, stratified by cognitive status. Mean task-evoked changes in phase noise standard deviation (ΔPnSD = post-task − pre-task) across harmonic orders (P1–P10) are shown for the Reference (Ref.) and mild cognitive impairment (MCI) groups. Points represent group means, and error bars indicate standard errors (SE).

**Fig 6.**
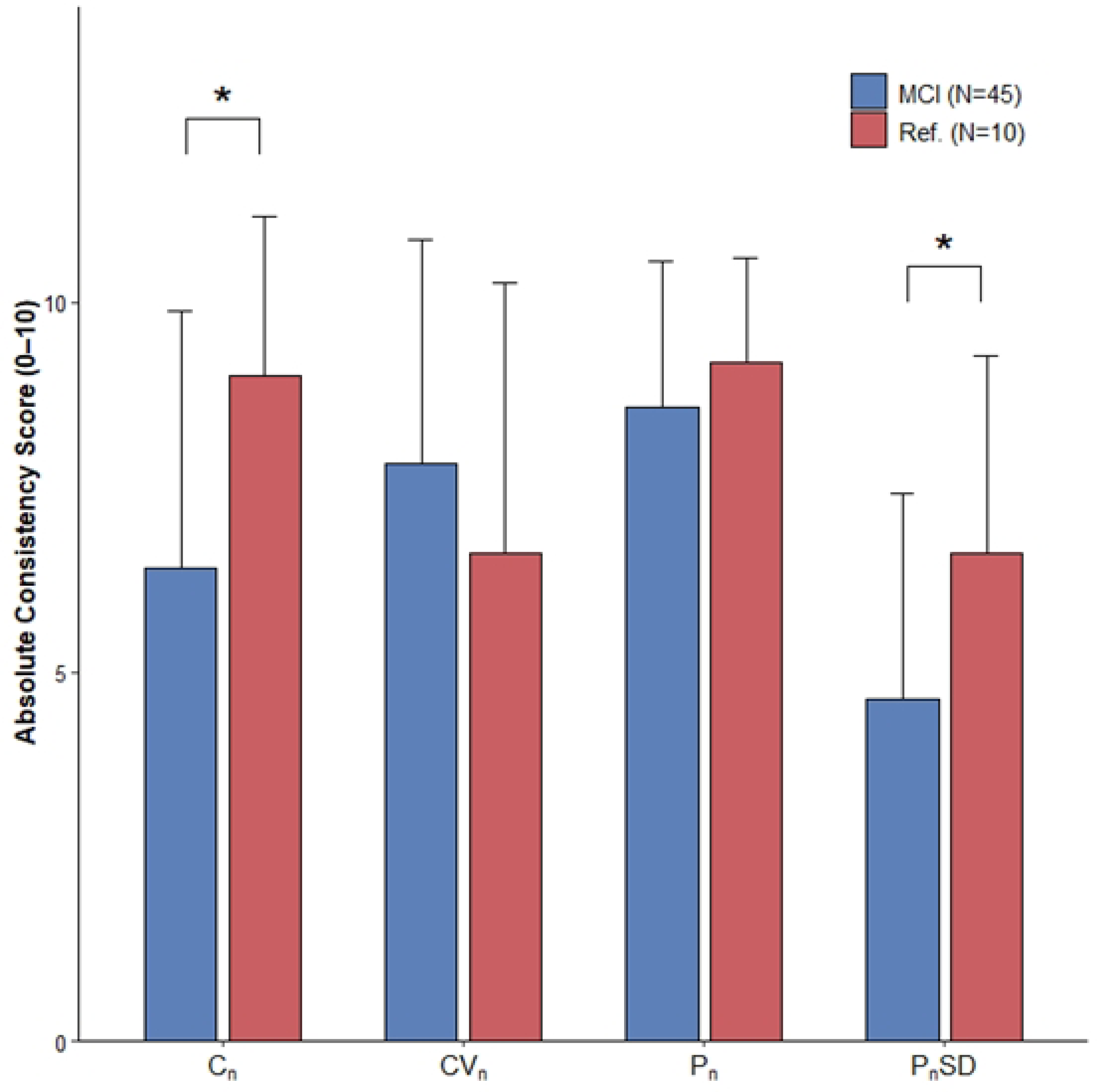
Comparison of absolute Harmonic Response Consistency Scores (HRCS) across physiological domains. Bar plots show mean absolute Harmonic Response Consistency Scores (HRCS) for harmonic amplitude (Cn), variability (CVn), phase angle (Pn), and phase noise (PnSD) domains in the Reference (Ref.; n = 10) and mild cognitive impairment (MCI; n = 45) groups. Error bars indicate standard deviations. Higher scores reflect greater directional coherence across harmonic orders. Asterisks indicate statistically significant between-group differences (*p < 0.05, Mann–Whitney U test).

### Associations Between Pulse Harmonics and Health-Related Measures

Pearson correlation analyses were conducted to examine associations between pulse harmonic parameters and cognitive/psychological measures (MoCA, AD8, BAI, BDI) and physiological measures (anthropometrics, blood pressure, and laboratory indices). Correlation analyses were performed using baseline pulse harmonic parameters. Across C1–C10, statistically significant associations were limited and concentrated in lower-order harmonics with systolic blood pressure. C1, C2, and C3 were negatively correlated with systolic blood pressure (C1: r = −0.218, p = 0.032; C2: r = −0.244, p = 0.017; C3: r = −0.250, p = 0.016). Other harmonics (C4–C10) showed no significant correlations with psychological, anthropometric, or metabolic markers.

For phase-related measures (P1–P10), a small number of low-magnitude associations were observed. P1 was positively correlated with BDI (r = 0.244, p = 0.018), and P6 was positively correlated with AD8 (r = 0.216, p = 0.037). Most other Pn correlations were weak and non-significant. For variability indices (CVn), correlations with clinical variables were generally small (|r| typically < 0.20) and not consistently significant. The correlation matrix is provided in S1 Table.

### Multidimensional Harmonic Response Consistency

The analysis of absolute Harmonic Response Consistency Scores (HRCS) revealed significant inter-group differences in physiological regulatory coherence. Specifically, for harmonic amplitude (|S_C_|), the Ref. group exhibited a remarkably high degree of consistency (Mean = 9.00 ± 2.16) compared to the significantly lower scores observed in the MCI group (Mean = 6.40 ± 3.48; Mann-Whitney U = 326.50, p = 0.020). This indicates that cognitively intact older adults maintained higher directional uniformity across harmonic orders in vascular tone across the frequency spectrum, whereas the MCI group displayed a fragmented response. A similar pattern was observed for phase noise consistency (|S_PnSD_|), where the Ref. group demonstrated significantly greater coherence than the MCI group (6.60 ± 2.68 vs. 4.62 ± 2.79; U = 322.50, p = 0.030), reflecting more synchronized modulation of temporal stability. As a robust check, independent-samples t-tests were also conducted and yielded consistent results for both amplitude and phase-noise coherence (Cn: p = 0.028; PnSD: p = 0.046). In contrast, no significant group differences were found for phase angle (|S_P_|) or variability (|S_CV_|) consistency (p > 0.05).

## Discussion

The present study examined pulse-wave harmonic characteristics as indicators of systemic physiological regulation in cognitive aging by jointly considering baseline associations and task-evoked responses within a single analytical framework. Two consistent observations emerged. First, cross-sectional analyses showed that baseline pulse harmonic features were only weakly associated with cognitive and psychological measures, with statistically significant correlations largely confined to lower-order harmonic amplitudes (C1–C3) and systolic blood pressure. This pattern indicates that resting pulse harmonics primarily reflect vascular-regulatory state rather than cognitive performance per se. Second, when the cardiovascular system was challenged by a standardized cognitive task, a qualitatively different pattern became evident: cognitively intact participants exhibited a directionally coherent down-modulation of the harmonics, whereas individuals in the MoCA-defined MCI stratum showed attenuated and directionally heterogeneous responses. Notably, these group differences were not captured by mean pre–post changes in individual parameters, but became apparent when response organization across harmonic orders was considered. Together, these findings indicate that pulse-harmonic features are more informative as markers of regulatory organization and adaptive coherence under cognitive demand than as static correlates of cognitive test scores.

Importantly, the observed findings support the view that cognitive decline in later life is accompanied by alterations in physiological regulation rather than by a single, disease-specific signal. Pulse harmonic parameters did not function as isolated markers directly corresponding to cognitive test scores or psychological indices. Instead, they reflected broader physiological states and response tendencies that became evident when examined across different phases of cognitive engagement [28]. Consistent with the exploratory aim of this study, the results emphasize physiological heterogeneity and regulatory diversity rather than diagnostic discrimination.

Cognitive engagement represents a transient physiological perturbation that challenges cardiovascular and autonomic regulation [29]. In this study, the subtraction task elicited subtle, pattern-level shifts in harmonic amplitudes and variability indices, characterized more by directionality and dispersion than by statistically significant mean changes. Among cognitively healthier participants, harmonic amplitudes tended to decrease following the cognitive task, suggesting a coordinated adjustment of cardiovascular dynamics in response to increased cognitive demand [30]. In contrast, participants with cognitive impairment showed smaller, inconsistent, or directionally heterogeneous changes.

The absence of statistically significant differences in mean pulse harmonic changes between groups should be interpreted in the context of physiological dysregulation. In cognitively healthier participants, the harmonic amplitudes tended to decrease after the subtraction task, a pattern consistent with a task-evoked shift in peripheral vascular regulation. Because concurrent autonomic or hemodynamic reactivity markers (e.g., heart rate, blood pressure reactivity, or HRV) were not recorded during the task, mechanistic interpretations such as sympatho-excitation–related vasoconstriction should be viewed as hypothesis-consistent rather than directly confirmed [31]. Some patients may exhibit paradoxical vasodilation while others show excessive vasoconstriction, leading to increased inter-individual variability that attenuates group-level mean effects.

The pronounced heterogeneity observed in the MCI group may be interpreted through the framework of "physiological complexity" [32] and "neurovisceral integration" [21, 33, 34], reflecting disrupted hierarchical autonomic-cognitive coupling and loss of fractal dynamics. According to the loss of complexity theory, healthy aging is characterized by the maintenance of adaptive, multi-scale regulatory networks [35, 36], whereas pathological states often exhibit a breakdown in these feedback loops, leading to either excessive rigidity or uncorrelated randomness [37, 38]. The findings suggest that the cardiovascular system in MCI participants does not simply fail to respond; rather, it responds incoherently. This phenomenon reflects a functional uncoupling between central cognitive demand and autonomic regulation [4]. When the precise synchronization between cortical activity and vascular tone is disrupted—potentially due to early autonomic neuropathy or endothelial dysfunction associated with cognitive decline [39]—the resulting pulse wave responses become directionally scattered [40]. Thus, the high inter-individual dispersion observed in the MCI group should not be viewed merely as statistical noise, but as a potential physiological marker of compromised systemic integration [41].

These patterns may be interpreted in terms of regulatory flexibility. Adaptive physiological systems are characterized by the capacity to adjust efficiently to contextual demands [42], whereas aging and cognitive decline have been associated with reduced responsiveness and diminished adaptability across multiple physiological domains [31, 43]. The attenuated or inconsistent task-evoked pulse responses observed in cognitively impaired participants may therefore reflect a reduced capacity for dynamic regulation rather than a simple absence of response [44, 45]. Notably, the lack of strong statistical significance across harmonic orders does not negate physiological relevance, as variability in response magnitude and direction is itself a salient feature of dysregulated systems [41, 46].

Beyond mean changes in harmonic amplitudes, variability indices provided additional insight into physiological regulation. CVn measures showed small and directionally mixed pre–post shifts, with larger dispersion in the cognitively impaired group. This suggests that cognitive engagement may amplify inter-individual variability rather than producing a uniform directional change.

From a physiological perspective, increased variability does not necessarily represent random noise [31]. In aging and disease contexts, excessive or poorly constrained variability has been interpreted as a marker of dysregulated physiological control rather than healthy complexity [35]. The greater dispersion of CVn observed in cognitively impaired participants may therefore reflect weakened coordination among cardiovascular regulatory mechanisms [47]. Lin, Hsiu (48) established that phase-angle variability reflects arterial transmission regulation, and the PnSD results exhibited similar directional tendencies. The lack of statistical significance suggests subtle systemic instability instead of clear structural problems, highlighting an important difference between these physiological conditions and the results shown by clinical scales.

Correlation analyses revealed that associations between pulse harmonic parameters and clinical or psychological measures were generally modest and confined to specific domains. Lower-order harmonic amplitudes (C1–C3) showed small but significant negative correlations with systolic blood pressure, consistent with established links between arterial properties and pulse waveform morphology [49]. In contrast, associations with cognitive scores (MoCA, AD8) and emotional measures (BAI, BDI) were sparse and weak.

Pulse harmonic features reflect integrated cardiovascular and autonomic dynamics rather than cognitive performance per se [17]. Cognitive test scores capture functional outcomes, whereas pulse harmonics represent underlying physiological states that underpin, but do not directly determine, cognitive function. The limited strength of these correlations underscores the conceptual distinction between physiological regulation and clinical assessment and supports the interpretation of pulse harmonics as contextual physiological indicators rather than surrogate cognitive measures.

Several methodological aspects strengthen the interpretability of the present findings. First, beat-level harmonic decomposition enabled detailed characterization of pulse waveform structure and variability across cardiac cycles, capturing both amplitude- and phase-related features [16]. Second, the within-subject pre–post task-evoked assessment allowed characterization of short-term physiological modulation associated with cognitive engagement, while reducing inter-individual confounding by focusing on within-person changes. This approach highlights context-dependent physiological responses without assuming uniform task effects across individuals [28]. Third, the community-based setting enhances ecological validity and demonstrates the feasibility of pulse-wave harmonic analysis outside specialized clinical environments.

Importantly, the study design was intentionally exploratory. Rather than testing a predefined diagnostic hypothesis, the analysis aimed to describe physiological patterns and response tendencies in a real-world aging population. This approach aligns with the complexity and heterogeneity inherent in cognitive decline and avoids oversimplification of multifactorial physiological processes.

Several limitations should be acknowledged. The sample exhibited imbalance in sex, education, and age across cognitive strata. Because participants were recruited from real-world community healthcare settings, the cohort reflects natural population composition rather than a demographically matched experimental sample. While this enhances ecological validity, it limits generalizability and introduces potential confounding effects, particularly by age and education. Notably, although the sample size for the paired Reference group is relatively small (n=10), the group exhibited high response consistency (mean HRCS = 9.00), which aligns with observations from previous studies in demographically matched samples. This suggests that the observed response patterns are physiologically robust rather than the result of random chance.

Cognitive engagement was assessed using a single subtraction task, which represents only one type of executive demand and may not capture the full spectrum of cognitive processes relevant to daily functioning. The cross-sectional design precludes inference regarding causal relationships or longitudinal trajectories of pulse harmonic dynamics; therefore, findings should be interpreted as descriptive patterns rather than confirmatory evidence.

Because age differed significantly across cognitive strata and blood pressure is age-sensitive in late life, baseline hemodynamic differences may partly reflect age-related vascular trajectories rather than cognitive status per se. Importantly, the primary findings concern task-evoked response organization (regulatory coherence) rather than baseline physiological levels, which may mitigate—but not eliminate—concerns related to demographic imbalance.

For phase-related measures (Pn), circular statistical approaches may be more appropriate than linear averaging; thus, these results should be interpreted cautiously. Correlation analyses were reported as unadjusted Pearson correlations in keeping with the exploratory aim, and residual confounding cannot be excluded.

The reference stratum was smaller and younger than the MCI group, which limits statistical power and may influence the magnitude and detectability of mean effects. However, this imbalance is less likely to fully account for the observed pattern-level findings. Specifically, the reference group consistently exhibited directionally coherent down-modulation of the harmonics and higher cross-harmonic consistency, features that reflect response organization rather than effect size. Notably, a highly similar harmonic response pattern was previously observed in an independent cohort explicitly matched for age, sex, and education [28], in which significant reductions in C2 and C3 and a concordant downward tendency in C1 were demonstrated following cognitive engagement. Taken together, these observations suggest that the present findings are unlikely to be solely driven by the age difference in the current community sample, although replication in larger and demographically balanced cohorts remains warranted.

Despite these limitations, the present study contributes to a growing body of evidence supporting a systemic perspective on cognitive decline [31, 50]. Pulse harmonic analysis offers a non-invasive means of characterizing cardiovascular and autonomic dynamics in community-dwelling older adults and may complement existing clinical and cognitive assessments [16]. Future studies incorporating longitudinal designs, multiple cognitive tasks, and multimodal physiological measures may further clarify how pulse harmonic patterns relate to trajectories of cognitive aging. Rather than serving as diagnostic biomarkers, pulse harmonic features may provide insight into physiological context, regulatory capacity, and inter-individual heterogeneity associated with cognitive decline. Such an approach may be particularly valuable for community-based research and population-level aging studies, where feasibility, non-invasiveness, and ecological validity are essential.

## Conclusion

In this community-based exploratory study, pulse-wave harmonic analysis revealed that resting harmonic features show only modest cross-sectional associations with cognitive and psychological measures, indicating limited value as static correlates of cognitive status. In contrast, cognitive stimulus exposed clear differences in regulatory organization. Cognitively intact older adults exhibited coherent down-modulation of low-order harmonics, whereas individuals in the MoCA-defined MCI stratum showed attenuated and directionally inconsistent responses. Importantly, these group differences were most robustly captured by coherence-based metrics, particularly the HRCS, rather than by mean pre–post changes in individual parameters. Collectively, these findings suggest that early cognitive decline is associated with disrupted physiological organization and reduced regulatory coherence, rather than a simple absence of cardiovascular responsiveness. Pulse-harmonic profiling may therefore serve as a non-invasive indicator of systemic regulatory capacity in community-dwelling older adults, complementing conventional cognitive assessments rather than functioning as a standalone diagnostic marker.

## Data Availability

All relevant data are within the manuscript and its Supporting Information files. The minimal data set, including de-identified demographic information, MoCA scores, and pulse-wave harmonic parameters, is provided as S1 Dataset.

## Acknowledgements

The author thanks the local community hubs and volunteers in the Cishan and Meinong areas for their assistance in study promotion and participant recruitment.

## Supporting information

**S1 Dataset. Minimal data set.** This file contains the de-identified underlying data for the study’s findings, including participants’ demographic information, MoCA scores, and pulse-wave harmonic parameters.

**S1 Table. Correlation matrix.** This table presents the Pearson correlation coefficients between pulse harmonic parameters and cognitive scores.

**S1 Checklist. STROBE checklist.** The reporting of this study conforms to the STROBE statement for observational studies.

## References

1. Kurakin A, Bredesen DE. Alzheimer’s disease as a systems network disorder: chronic stress/dyshomeostasis, innate immunity, and genetics. Aging (Albany NY). 2020;12(18):17815.

2. Morris JK, Honea RA, Vidoni ED, Swerdlow RH, Burns JM. Is Alzheimer’s disease a systemic disease? Biochimica et Biophysica Acta (BBA)- Molecular Basis of Disease. 2014;1842(9):1340–9.

3. Cortes CJ, Thyfault JP, Wilkins HM. Editorial: Systemic implications of Alzheimer’s disease. Frontiers in Aging Neuroscience. 2023;Volume 15 - 2023. doi: 10.3389/fnagi.2023.1219987.

4. Forte G, Favieri F, Casagrande M. Heart rate variability and cognitive function: A systematic review. Frontiers in neuroscience. 2019;13:710.

5. Pugazhenthi S, Qin L, Reddy PH. Common neurodegenerative pathways in obesity, diabetes, and Alzheimer’s disease. Biochimica et biophysica acta (BBA)-molecular basis of disease. 2017;1863(5):1037–45.

6. Sweeney MD, Montagne A, Sagare AP, Nation DA, Schneider LS, Chui HC, et al. Vascular dysfunction—the disregarded partner of Alzheimer’s disease. Alzheimer’s & Dementia. 2019;15(1):158–67.

7. Boah M, Cyuzuzo C, Uwinkindi F, Kalinda C, Yohannes T, Greig C, et al. Frailty, multimorbidity and quality of life in an ageing population in Africa: a cross-sectional, population-based study in rural and urban Rwanda. Family Medicine and Community Health. 2025;13(4):e003512.

8. Koutsonida M, Markozannes G, Bouras E, Aretouli E, Tsilidis KK. Metabolic syndrome and cognition: A systematic review across cognitive domains and a bibliometric analysis. Frontiers in Psychology. 2022;13:981379.

9. Zuo Q, Song L, Gao X, Cen M, Fu X, Qin S, et al. Associations of metabolic syndrome with cognitive function and dementia risk: Evidence from the UK Biobank cohort. Diabetes, Obesity and Metabolism. 2024;26(12):6023–33.

10. Sipilä K, Koivistoinen T, Moilanen L, Nieminen T, Reunanen A, Jula A, et al. Metabolic syndrome and arterial stiffness: the Health 2000 Survey. Metabolism. 2007;56(3):320–6.

11. Ejiri K, Ding N, Kim E, Honda Y, Cainzos-Achirica M, Tanaka H, et al. Association of Segment-Specific Pulse Wave Velocity With Vascular Calcification: The ARIC (Atherosclerosis Risk in Communities) Study. Journal of the American Heart Association. 2024;13(2):e031778.

12. Pereira T, Paulino E, Maximiano S, Rosa M, Pinto AL, Mendes MJ, et al. Measurement of arterial stiffness and vascular aging in community pharmacies—The ASINPHAR@ 2action project. The Journal of Clinical Hypertension. 2019;21(6):813–21.

13. Wu H-T, Haryadi B, Chen J-J. A first step towards a comprehensive approach to harmonic analysis of synchronous peripheral volume pulses: a proof-of-concept study. Journal of Personalized Medicine. 2021;11(12):1263.

14. Wu H-T, Wu H-K, Wang C-L, Yang Y-L, Wu W-H, Tsai T-H, et al. Modeling the pulse signal by wave-shape function and analyzing by synchrosqueezing transform. PloS one. 2016;11(6):e0157135.

15. Bae J-H, Jeon YJ. Pulse sharpness as a quantitative index of vascular aging. Scientific Reports. 2021;11(1):19895.

16. Chang C-W, Chen J-M, Wang W-K. Development of a standard protocol for the harmonic analysis of radial pulse wave and assessing its reliability in healthy humans. IEEE J Transl Eng Health Med. 2015;3:1–6.

17. Hsiao K-Y, Kang J-H, Wu Y-S, Chang H-H, Yang C-T. Peripheral pulse harmonic analysis and its clinical application: A systematic review. Journal of Traditional and Complementary Medicine. 2024;14(2):223–36.

18. Ayres P, Lee JY, Paas F, Van Merrienboer JJ. The validity of physiological measures to identify differences in intrinsic cognitive load. Frontiers in psychology. 2021;12:702538.

19. Pavlov YG, Gashkova AS, Kasanov D, Kosachenko AI, Kotyusov AI, Kotchoubey B. Task-evoked pulse wave amplitude tracks cognitive load. Scientific Reports. 2023;13(1):22592.

20. Thayer JF, Mather M, Koenig J. Stress and aging: A neurovisceral integration perspective. Psychophysiology. 2021;58(7):e13804.

21. Condy EE, Friedman BH, Gandjbakhche A. Probing neurovisceral integration via functional near-infrared spectroscopy and heart rate variability. Frontiers in Neuroscience. 2020;14:575589.

22. Oppenheimer S, Cechetto D. The insular cortex and the regulation of cardiac function. Comprehensive Physiology. 2016;6(2):1081–133.

23. Nicolini P, Ciulla MM, Malfatto G, Abbate C, Mari D, Rossi PD, et al. Autonomic dysfunction in mild cognitive impairment: evidence from power spectral analysis of heart rate variability in a cross-sectional case-control study. PloS one. 2014;9(5):e96656.

24. Nicolini P, Lucchi T, Abbate C, Inglese S, Tomasini E, Mari D, et al. Autonomic function predicts cognitive decline in mild cognitive impairment: Evidence from power spectral analysis of heart rate variability in a longitudinal study. Frontiers in aging neuroscience. 2022;14:886023.

25. Issac TG, Chandra SR, Gupta N, Rukmani MR, Deepika S, Sathyaprabha T. Autonomic dysfunction: A comparative study of patients with Alzheimer’s and frontotemporal dementia–A pilot study. Journal of neurosciences in rural practice. 2017;8(1):84.

26. Chou Y-T, Sun Z-J, Shao S-C, Yang Y-C, Lu F-H, Chang C-J, et al. Autonomic modulation and the risk of dementia in a middle-aged cohort: A 17-year follow-up study. biomedical journal. 2023;46(6):100576.

27. Mizukami K. Autonomic dysfunction in dementia with Lewy bodies: Focusing on cardiovascular and respiratory dysfunction. Psychiatry and Clinical Neurosciences Reports. 2023;2(3):e129.

28. Hsiao K-Y. Theoretical Validation of Pulse Wave Harmonic Features and Their Alignment with Traditional Chinese Medicine Theory in Response to Cognitive Stimulation [Doctoral dissertation]. Tainan, Taiwan: Institute of Health Care Science, National Cheng Kung University; 2025.

29. Stenfors CU, Hanson LM, Theorell T, Osika WS. Executive cognitive functioning and cardiovascular autonomic regulation in a population-based sample of working adults. Frontiers in Psychology. 2016;7:1536.

30. Liebel SW, Jones EC, Oshri A, Hallowell ES, Jerskey BA, Gunstad J, et al. Cognitive processing speed mediates the effects of cardiovascular disease on executive functioning. Neuropsychology. 2017;31(1):44.

31. Manor B, Lipsitz LA. Physiologic complexity and aging: implications for physical function and rehabilitation. Progress in Neuro-Psychopharmacology and Biological Psychiatry. 2013;45:287–93.

32. Das S, Puthankattil SD. Functional connectivity and complexity in the phenomenological model of mild cognitive-impaired Alzheimer’s disease. Frontiers in Computational Neuroscience. 2022;16:877912.

33. Thayer JF, Lane RD. Claude Bernard and the heart–brain connection: Further elaboration of a model of neurovisceral integration. Neuroscience & Biobehavioral Reviews. 2009;33(2):81–8.

34. de Oliveira Matos F, Vido A, Garcia WF, Lopes WA, Pereira A. A neurovisceral integrative study on cognition, heart rate variability, and fitness in the elderly. Frontiers in aging neuroscience. 2020;12:51.

35. Goldberger AL, Amaral LA, Hausdorff JM, Ivanov PC, Peng C-K, Stanley HE. Fractal dynamics in physiology: alterations with disease and aging. Proceedings of the national academy of sciences. 2002;99(suppl_1):2466–72.

36. Costa M, Goldberger AL, Peng CK. Multiscale Entropy Analysis of Complex Physiologic Time Series. Physical Review Letters. 2002;89(6):068102. doi: 10.1103/PhysRevLett.89.068102.

37. Lipsitz LA, Goldberger AL. Loss of’complexity’and aging: potential applications of fractals and chaos theory to senescence. Jama. 1992;267(13):1806–9.

38. Sleimen-Malkoun R, Temprado J-J, Hong SL. Aging induced loss of complexity and dedifferentiation: consequences for coordination dynamics within and between brain, muscular and behavioral levels. Frontiers in Aging Neuroscience. 2014;6:140.

39. Toth P, Tarantini S, Csiszar A, Ungvari Z. Functional vascular contributions to cognitive impairment and dementia: mechanisms and consequences of cerebral autoregulatory dysfunction, endothelial impairment, and neurovascular uncoupling in aging. American Journal of Physiology-Heart and Circulatory Physiology. 2017;312(1):H1–H20.

40. You T-Y, Dong Q, Cui M. Emerging links between cerebral blood flow regulation and cognitive decline: a role for brain microvascular pericytes. Aging and Disease. 2023;14(4):1276.

41. McCraty R, Shaffer F. Heart rate variability: new perspectives on physiological mechanisms, assessment of self-regulatory capacity, and health risk. Global advances in health and medicine. 2015;4(1):46–61.

42. Pomatto LC, Davies KJ. The role of declining adaptive homeostasis in ageing. The Journal of physiology. 2017;595(24):7275–309.

43. Arbeev KG, Ukraintseva SV, Bagley O, Zhbannikov IY, Cohen AA, Kulminski AM, et al. “Physiological Dysregulation” as a promising measure of robustness and resilience in studies of aging and a new indicator of preclinical disease. The Journals of Gerontology: Series A. 2019;74(4):462–8.

44. Gao C, Lim AS, Haghayegh S, Cai R, Yang J, Yu L, et al. Reduced Complexity of Pulse Rate Is Associated With Faster Cognitive Decline in Older Adults. Journal of the American Heart Association. 2025;14(10):e041448.

45. Thorin-Trescases N, de Montgolfier O, Pinçon A, Raignault A, Caland L, Labbé P, et al. Impact of pulse pressure on cerebrovascular events leading to age-related cognitive decline. American Journal of Physiology-Heart and Circulatory Physiology. 2018;314(6):H1214–H24.

46. Saul JP, Valenza G. Heart rate variability and the dawn of complex physiological signal analysis: methodological and clinical perspectives. Philosophical Transactions of the Royal Society A: Mathematical, Physical and Engineering Sciences. 2021;379(2212).

47. Kuo Y-C, Chiu T-Y, Jan M-Y, Bau J-G, Li S-P, Wang W-K, et al. Losing harmonic stability of arterial pulse in terminally ill patients. Blood pressure monitoring. 2004;9(5):255–8.

48. Lin F-C, Hsiu H, Chiu H-S, Chen C-T, Hsu C-H. Characteristics of pulse-waveform and laser-Doppler indices in frozen-shoulder patients. Biomedical Signal Processing and Control. 2020;56:101718.

49. Nichols WW, Denardo SJ, Wilkinson IB, McEniery CM, Cockcroft J, O’Rourke MF. Effects of arterial stiffness, pulse wave velocity, and wave reflections on the central aortic pressure waveform. The journal of clinical hypertension. 2008;10(4):295–303.

50. Jost Z, Kujach S. Understanding Cognitive Decline in Aging: Mechanisms and Mitigation Strategies–A Narrative Review. Clinical Interventions in Aging. 2025:459–69.

